# Modelling the SARS-CoV-2 first epidemic wave in Greece: social contact patterns for impact assessment and an exit strategy from social distancing measures

**DOI:** 10.1101/2020.05.27.20114017

**Authors:** Vana Sypsa, Sotirios Roussos, Dimitrios Paraskevis, Theodore Lytras, S Sotirios Tsiodras, Angelos Hatzakis

**Affiliations:** Department of Hygiene, Epidemiology and Medical Statistics, Medical School, National and Kapodistrian University of Athens, Athens, Greece; National Public Health Organization, Athens, Greece; 4^th^ Department of Internal Medicine, Medical School, National and Kapodistrian University of Athens, Athens, Greece

## Abstract

In Greece, a nationwide lockdown to mitigate the transmission of SARS-CoV-2 was imposed on March 23, 2020. As by the end of April the first epidemic wave is waning, it is important to assess the infection attack rate and quantify the impact of physical distancing. We implemented a survey to assess social mixing patterns before the epidemic and during lockdown. We estimated R_0_ from surveillance data and assessed its decline as a result of physical distancing based on social contacts data. We applied a Susceptible-Exposed-Infectious-Recovered model to estimate the infection attack rate and the infection fatality ratio (IFR). As multiple social distancing measures were implemented simultaneously (schools/work/leisure), we assessed their overall impact as well as their relative contribution. R_0_ was estimated 2·38 (95%CI: 2·01,2·80). By April 26^th^, the infection attack rate was 0·12% (95%CrI: 0·06%,0·26%) and the IFR 1·12% (95%CrI: 0·55%,2·31%). During lockdown, daily contacts were reduced by 86·9% and the effective reproduction number reached 0·46 (95%CrI: 0·35,0·57). The reduction in R_0_ attributed to lockdown was 81·0% (95%CrI: 71·8%,86·0%) whereas the reduction attributed to each measure separately ranged between 10%-24%. We assessed scenarios with less disruptive social distancing measures as well as scenarios where measures are partially lifted after lockdown. This is the first impact assessment of the first wave of SARS-CoV-2 in a European country. It suggests that only multiple measures implemented simultaneously could reduce R_0_ below 1. Measuring social mixing patterns can be a tool for real-time monitoring of the epidemic potential.

## Introduction

Coronavirus disease 2019 (COVID-2019) caused by a novel coronavirus [severe acute respiratory syndrome coronavirus-2 (SARS-CoV-2)] emerged in Wuhan, China in December 2019 [1] and spread worldwide in 212 countries and territories causing more than 2,8 million cases and 194,000 deaths within a period of 4 months. [2] The transmission potential of SARS-Cov-2 is high as a consequence of the high basic reproduction number (R_0_) and the lack of previous immunity in humans.[3, 4] Fatality rates of COVID-19 are higher in older populations and low-income countries with underdeveloped hospital infrastructure and critical care facilities. [5, 6]

The goal of public health authorities is the strongest possible mitigation in order to suppress the epidemic curve until sufficient population immunity and/or availability of effective therapies/vaccine. Central strategies include travel-related and a variety of case-based and social distancing interventions. Social distancing or community measures aim to decrease social contacts and contain the transmission. [7]

In Greece, the first case was reported on February 26, 2020 and, soon after, a number of social distancing in addition to travel related and case-based interventions were implemented. A nationwide lockdown took place on March 23 (Figure 1) (SI Appendix, Table S1). By the end of April, the first wave of the epidemic has waned and withdrawal of physical distancing interventions is a social priority. Greece has been referred as an example of a country with successful response against COVID-19, despite the severe financial crisis experienced in the recent years and its aged population. [8] However, given the substantial risk of re-emergence of a second wave, careful consideration and close monitoring is needed to inform strategies on how to withdraw social distancing and resume social and economic activities.

**Figure 1.**
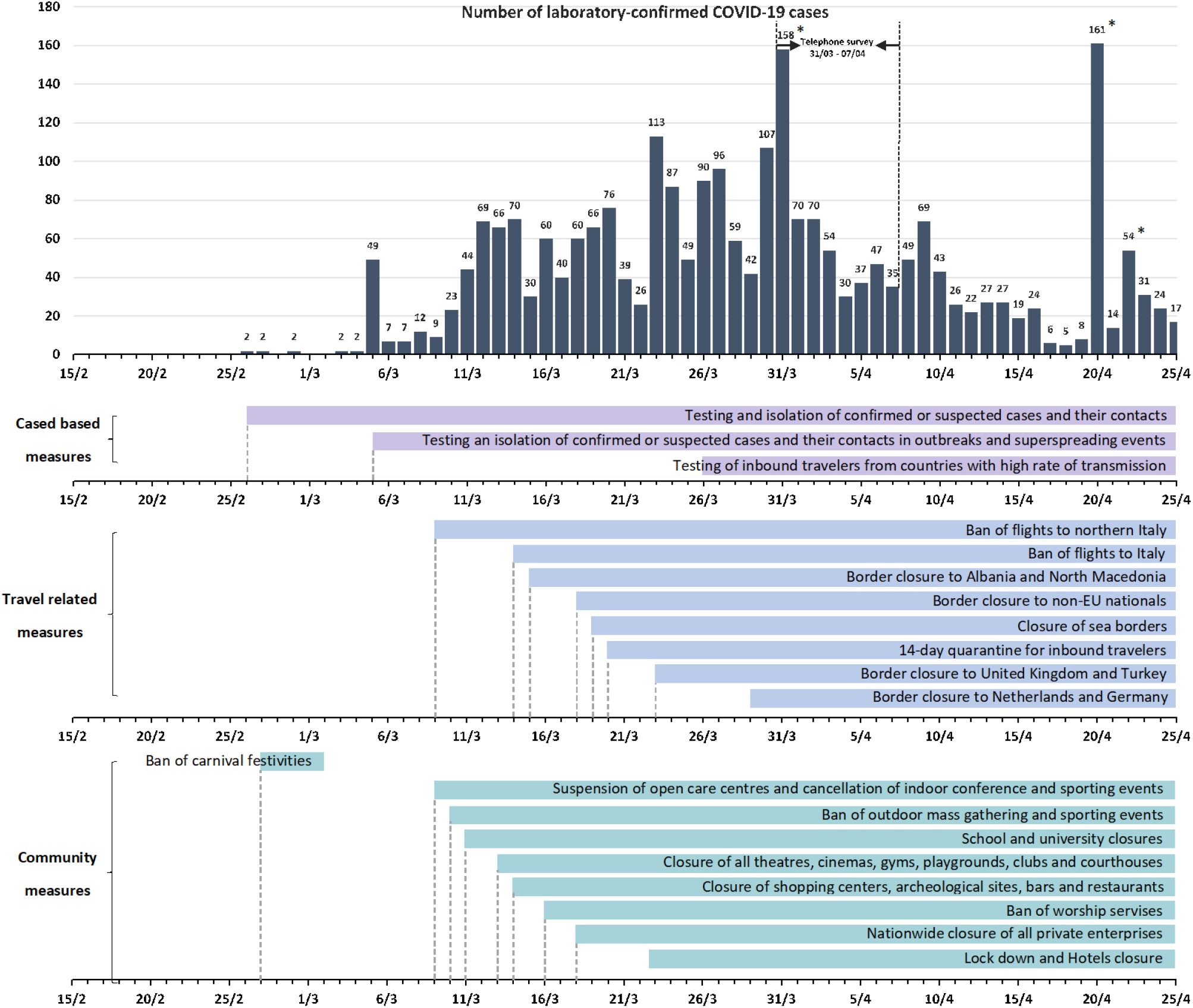
Daily number of cases by date of sampling for laboratory testing [11] and timeline of key measures. The high number of diagnosed cases at the end of March as well as in late April (bars indicated with an asterisk) correspond to clusters of cases in three settings (a ship, a refugee camp and a clinic)

Previous impact assessments of the first epidemic wave were published for Wuhan and major Chinese cities. [7, 9, 10] In this paper, we provide estimates for the first wave of the epidemic in Greece based on modelling and report an assessment of the effect of social distancing measures. This is the first impact assessment of the first wave in a European country.

## Results

### The course of the epidemic in Greece (15 February-26 April 2020)

The number of confirmed cases by date of sampling for laboratory testing and the timeline of the various social distancing measures in Greece are depicted in Figure 1. By April 26^th^, 2020, there were 2,517 diagnosed cases (23·0% were imported) and 134 deaths [11]. The corresponding naive case fatality rate (CFR) in Greece is 5·3%.

### Model predictions

R_0_ was estimated 2·38 (95% CI: 2·01, 2·80). The course of R_t_ over time is shown in Figure 2A. The initial decline resulted from the fist social distancing measures (closure of schools, shops, restaurants etc.) and the second drop from the implementation of restrictions of all non-essential movement throughout the country (lockdown). The estimated R_t_ during lockdown is 0·46 (95% CrI: 0·35, 0·57).

**Figure 2.**
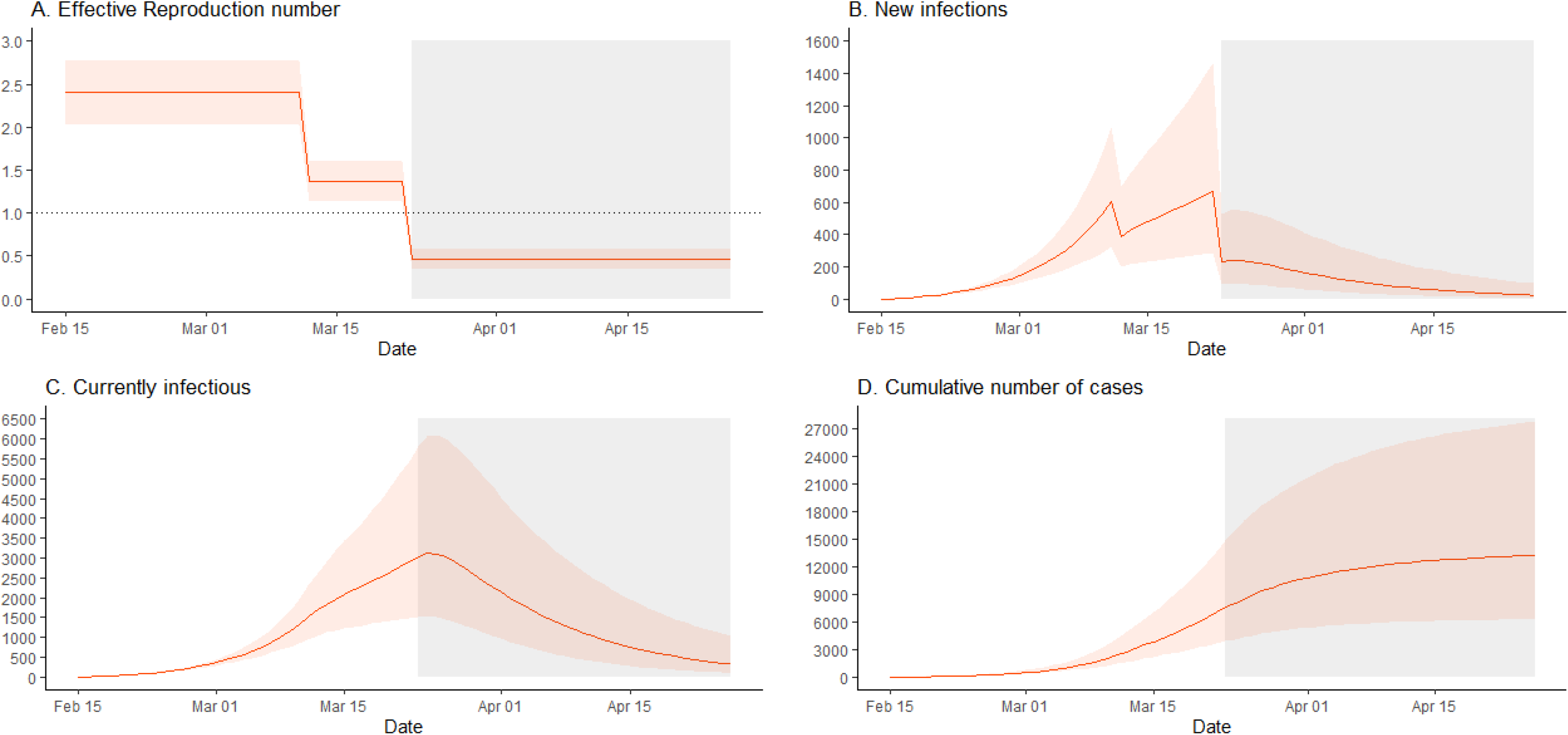
The first wave of the SARS-CoV-2 epidemic in Greece (15 February-26 April 2020) The grey rectangle indicates the period of restrictions of all non-essential movement in the country (lockdown).

At the end of the simulations period (April 26^th^), it is estimated that there are 25 new infections per day (95% CrI: 6, 97) and 329 infectious cases (95% CrI: 97, 1027) (Figures 2B, 2C). The infection attack rate is 0·12% (95% CrI: 0·06%, 0·26%). This corresponds to 13,189 infections in total since the start of the epidemic (95% CrI: 6,206, 27,700) (Figure 2D). The case ascertainment is estimated 19·1% (95% CrI: 9·1%, 40·6%).

Based on the number of deaths and critically ill patients reported by April 26^th^ and using as denominator the number of infections obtained from the model with the appropriate time lag, the IFR and the cumulative proportion of critically ill patients in Greece is 1·12% (95% CrI: 0·55%, 2·31%) and 1·55% (95% CrI: 0·75%, 3·22%), respectively.

To validate the model, we compared the reported number of deaths to that estimated by applying an age-adjusted published estimate of IFR to the number of infections predicted by the model. By April 26^th^, the reported number of deaths was 134; the predicted number of deaths is 137 (95% CrI: 66, 279). The predicted daily number of deaths versus the observed is shown in Figure S1.

As a sensitivity analysis, we simulated the epidemic and calculated these quantities assuming a shorter serial interval, finding very similar results concerning the infection attack rate and the IFR (Figure S2).

### Disentangling the relative contribution of social distancing measures

In total, 602 persons provided contact diaries; 12,463 and 1,743 contacts were reported in the periods before the pandemic and during lockdown, respectively (Table 1). The mean daily number of contacts declined from 20·7 to 2·9 for the lockdown period (adjusted for the age distribution of the Greek population: 19·9 vs 2·6, reduction: 86·9%).

**Table 1.**
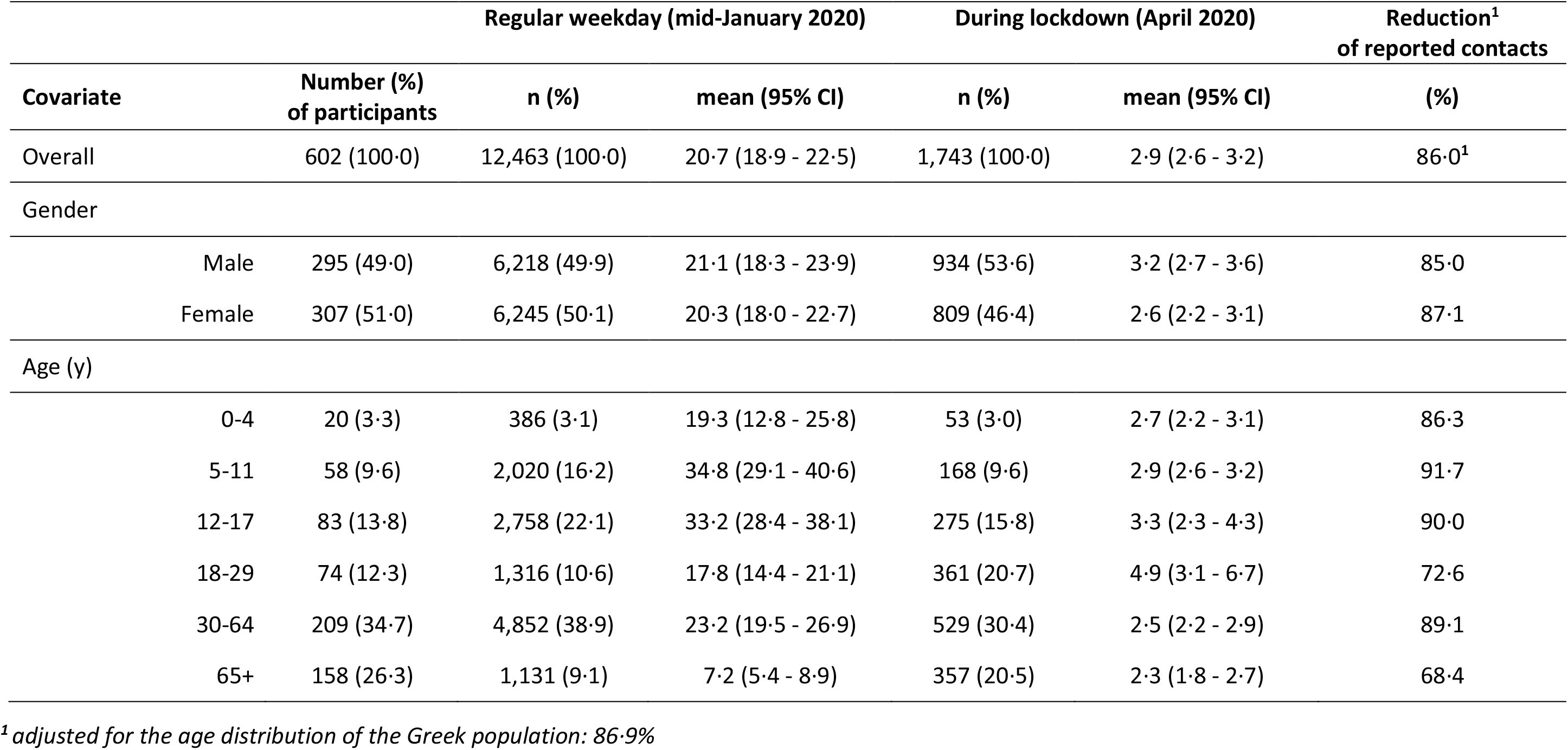
Number of contacts on a weekday during lockdown (March 31-April 7, 2020) and on the corresponding day before the epidemic (January 2020) in Athens, Greece (N=602 participants)

The change in age-mixing patterns is shown in the contact matrices of Figure 3A. In the pre-pandemic period, the diagonal of the contact matrix depicts the assortativity by age as participants tend to associate more often with other people of similar age. When social distancing measures were put into effect, the assortativity by age disappeared and contacts occurred mainly between household members (Figures 3B-3D).

**Figure 3.**
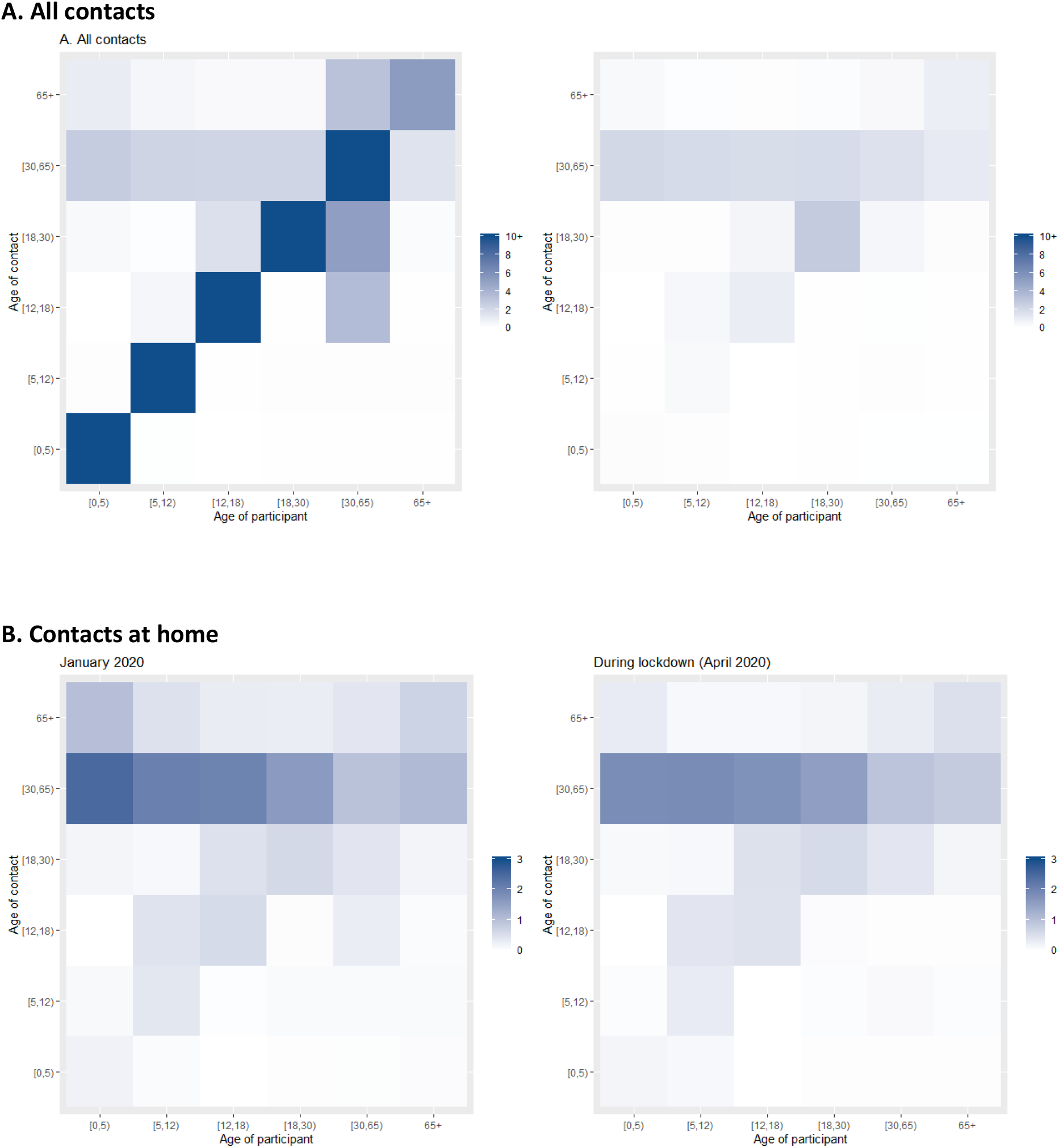
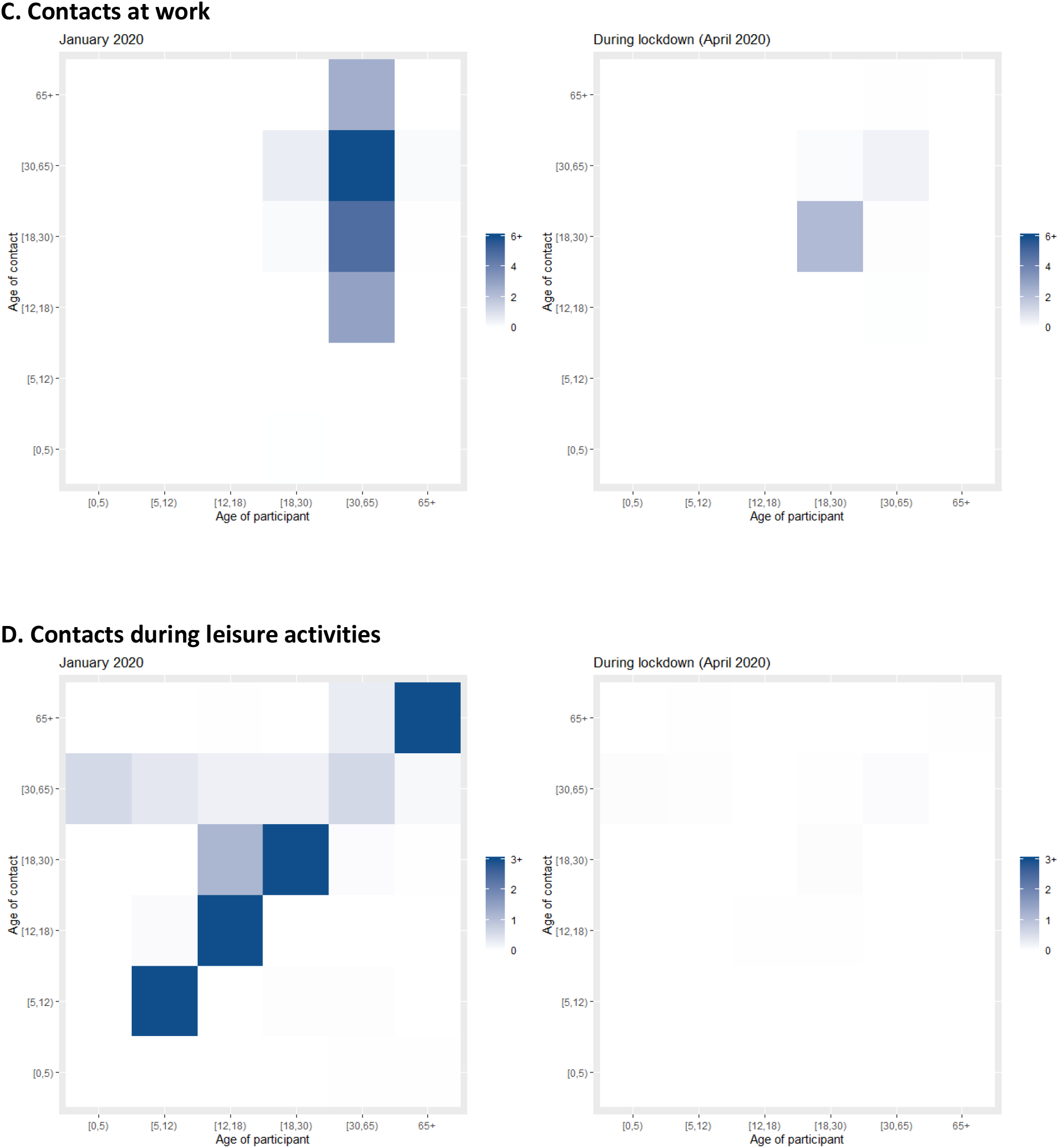
Age-specific contact matrices in Greece before the pandemic (January 2020) and during lockdown (April 2020) Each cell represents the average number of reported contacts, stratified by the age group of the participants and their corresponding contacts. In Figure 3A (all contacts), the diagonal of the contact matrix corresponds to contacts between people of the same age group, the bottom left corner of the matrix corresponds to contacts between school age children and the central part to contacts mainly at the work environment.

Based on the age-specific contact matrices of the two periods, the estimated reduction of R_0_ resulting from lockdown is 81·0% (95% CrI: 71·7%, 86·1%) (Figure 4). Thus, lockdown would have reduced R_t_ to below 1·0 even if the initial R_0_ had been as high as 5·3 (95% CrI: 3·5, 7·2).

**Figure 4.**
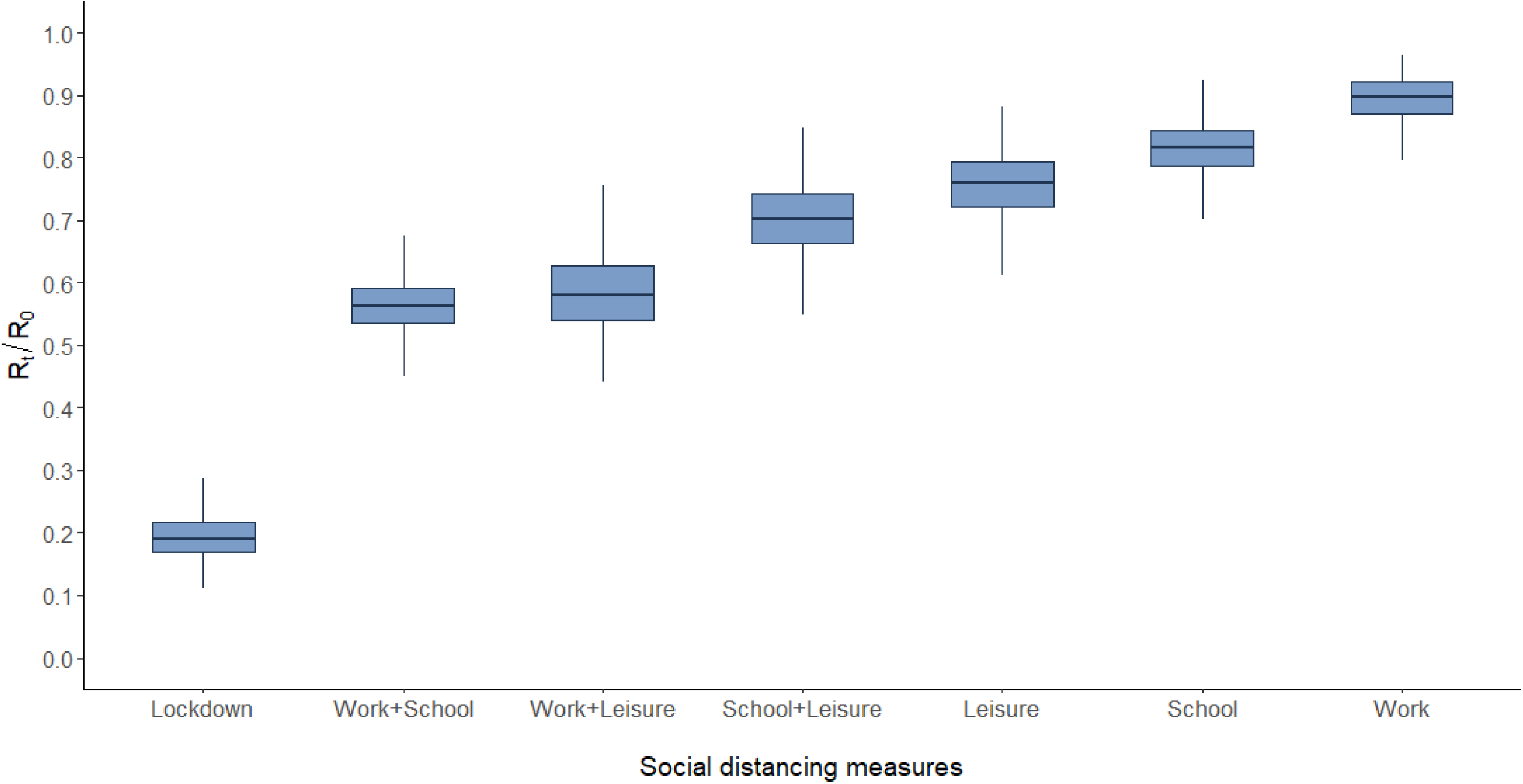
Estimated ratio R_t_/R_0_ to assess the impact of lockdown as well as the relative contribution of each measure or combination of measures as implemented during lockdown in Greece. A ratio of e.g. 0·80 corresponds to 20% decline in R_0_. The lowest ratio, corresponding to the period of lockdown, was obtained by comparing the social contacts data collected for the two periods (April 2020 vs January 2020). The other estimates were derived using the information from contact diaries in January 2020 corresponding to a regular school/working day and excluding or reducing subsets of social contacts at school, work home and leisure, based on what was observed during lockdown.

The impact of each measure separately as well as of combinations of measures is shown in Figure 4. The reduction in R_0_ attributed to each one of these measures is estimated 18·5% (95% CI: 10·7%,26·3%) for school closure, 10·3% (95% CI: 5·2%, 20·3%) for the decline in work contacts and 24·1% (95% CI: 14·8%, 34·3%) for the decline in contacts in leisure activities. The application of each measure separately would have reduced R_t_ to below 1·0 if the initial R_0_ had been as high as 1·23 (schools), 1·11 (work) and 1·32 (leisure). Combination of measures could be effective for higher R_0_ (work and school: 1·78, work and leisure: 1·72, school and leisure: 1·43).

We assessed alternative scenarios with less disruptive social distancing measures. For example, a reduction of 50% in school contacts (e.g. smaller classrooms), 20% in work contacts (e.g. teleworking) and 20% in leisure activities could reduce R_t_ to below 1·0 for initial levels as high as 1·32 (95% CrI: 1·27, 1·38). An even higher decline in leisure activities (50%), could be successful for an initial R_0_ as high as 1·48 (95% CrI: 1·35, 1·62).

Finally, we estimated the increase in R_t_ anticipated following the partial lift of lockdown measures assuming that contacts at work, school and leisure activities will return to levels that are 50%, 50% and 60% lower compared to pre-epidemic levels, respectively, and that the implementation of infection control measures (hand hygiene, masks, keeping distances) may reduce susceptibility to infection by 5%-30%. R_t_ will remain below 1 assuming at least 20% reduction in susceptibility as a result of these measures (Figure 5).

**Figure 5.**
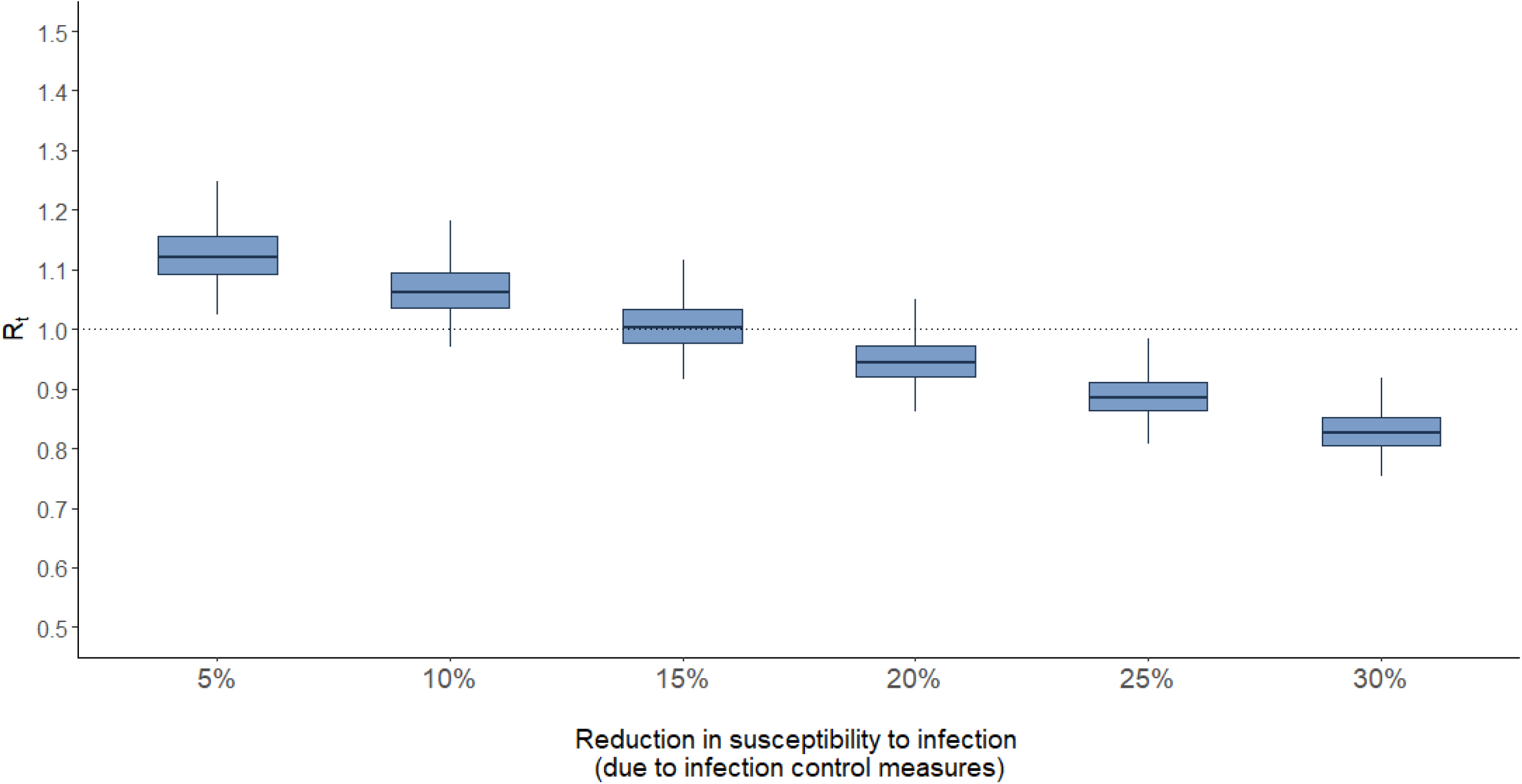
Estimated R_t_ following the partial lift of social distancing measures at the end of the first SARS-CoV-2 wave in Greece depending on the efficacy and adherence to infection control measures (hand hygiene, use of masks, keeping distances) R_t_ during lockdown was 0·46. For the partial lift of measures, we hypothesised a scenario where contacts at work, school and leisure activities will return to levels that are 50%, 50% and 60% lower compared to pre-epidemic levels, respectively.

## Discussion

As the first wave of the SARS-CoV-2 epidemic is waning in many countries, it is important to assess the burden of infection and of death in the population and to quantify the impact of social distancing. First, because these stringent measures have a major economic impact and restrict individual freedom. Second, lockdown leads to low infection attack rates in many countries raising the possibility of a second wave following the resumption of some of the economic activities and travel; thus, some type of location-specific physical distancing measures may need to be re-implemented in the near future.

As of May 18^th^, 2020, Greece has one of the lowest number of reported COVID-19 deaths in Europe (15·2 deaths per million population) [2] (SI Appendix, Table S2). Accordingly, our estimate of the infection attack rate was very low (0·12%). To simulate the outbreak in Greece, we estimated R_0_, assessed its decline over time, as a result of social distancing measures, and applied those in a mathematical model to estimate infections and deaths. Other research groups have applied back calculation of infections from reported deaths in a Bayesian framework. [12, 13] The resulting infection attack rate under the two approaches was almost identical (0·12% vs. 0.13%). [13] This attack rate is one of the lowest in Europe [13] and reflects the substantial impact of preventive measures in mitigating transmission. A consequence of this low attack rate is the possibility of continuing transmission if measures are lifted, as this is the case even for countries that were more severely hit. According to our estimates, the number of infectious cases has subsided considerably towards the end of April and social distancing measures can be lifted gradually. However, even in this period with low transmission levels, two local outbreaks were identified, the first in a refugee camp and the second in a private health care unit, thus increasing the number of diagnosed cases in the respective days (Figure 1). An increasing number of reports around the world suggest the significance of super spreading events [14-17] and caution should be exercised to prevent them or recognize them early.

In the validation of the model, the resulting number of deaths (137 (95% CrI: 66, 279) was in agreement with the observed (134 deaths). In addition, the estimated IFR of 1·12% was similar to that anticipated for the Greek population based on a published estimate, adjusted for non-homogenous attack rates by age and for demography. [5] The IFR of 0·66% estimated for mainland China [5] is 1·14% after standardizing for the age distribution of the Greek population.

We assessed the impact of social distancing by directly measuring individuals’ contact patterns during lockdown using a social contacts survey in a sample including children. Only two other social contacts surveys have been implemented during COVID-19 lockdown (in China and UK) and only one of those included children. [18, 19] These studies had common findings: there was a large reduction in the number of contacts (86·9% in Greece, 90·3% in Shanghai, 86·4% in Wuhan and 73·1% in UK; the latter refers to adults only), the assortativity by age – i.e. contacts between people of the same age group - disappeared and contacts during that period were mainly among household members. Other studies have assessed the impact of social distancing indirectly using social contacts data from pre-epidemic periods and assuming that interventions reduce social mixing in different contexts. [20, 21]

We estimated that R_0_ declined by 81·0% and reached 0·46 during lockdown. This is in agreement with estimates from China [7, 22] as well as from UK (76·2%) and France (84%). [23, 24] In our analysis, we assumed lower susceptibility to infection among children as there is growing body of evidence supporting this. [18, 25-28]

We further attempted to disentangle the impact of each measure. School closures, for example, were deployed early in China, Hong Kong, Singapore and many countries had instituted large-scale or national closure of schools, including Greece. [29] Based on our findings, only lockdown may reduce R_t_ below 1·0 for an initial R_0_ at the levels observed in Greece (around 2·4). Each measure - when implemented at the levels observed during lockdown in Greece - can reduce R_t_ to levels below 1·0 for initial R_0_ around 1·2. We should note that there is an interrelation between the different measures and the approach we have used might be an approximation; for example, school closure might result in increases in leisure contacts or decline in work contacts as parents need to attend younger children. However, it provides some information about the individual impact of each measure. Combinations of measures, such as reductions in contacts at work and school, would be successful for higher initial R_0_ near 1·8. We also assessed a scenario with less disruptive social distancing measures that could reduce R_t_ below 1·0 for initial levels as high as 1·33. Concerning the course of the epidemic post-lockdown, the effectiveness of infection control measures in reducing transmission is not known. We considered scenarios with levels varying between 5%-30%. It seems that an effectiveness of infection control measures higher than 15% in combination with less disruptive social distancing may keep R_t_ below 1·0.

Our analysis has some limitations. First, the lack of baseline pre-epidemic data since this was the first social contacts survey implemented in Greece. We asked respondents to report their contacts approximately two months earlier than the survey to make sure that these contacts were not affected by increased awareness of the pandemic. There might be recall bias although it is not clear to what direction. Our analysis has focused on the number of contacts, their ages (using wide age groups) and the place of the contact and not in the duration or the type of contact (physical or not) where the bias might be higher. Second, the survey was conducted in a sample from Athens Metropolitan Area and not from the whole of Greece. However, no relationship has been found between social contacts and urbanization. [30] In addition, the majority of the population lives in urban areas (Athens and other cities account for 35% and 79% of the Greek population, respectively) and the observed reduction of social contacts during lockdown was similar to other surveys conducted in China and UK.[24, 31] Third, 7the estimate of R_0_ depends on the serial interval and the number of imported cases. As there was no data from a local study of pairs infector-infectee, the distribution of the serial interval was based on previous estimates.[12, 32-34] The estimated R_0_ was in accordance with the epidemics in China [4] and Italy [35] and in our simulations we accounted for the uncertainty in this value. In addition, we repeated the analysis assuming a shorter serial interval [36]; this resulted in a lower R_0_ and R_t_ (i.e. a more optimistic scenario in terms of epidemic control through social distancing). Fourth, the environmental component of R_0_ (fomites etc.) was not considered due to the lack of data. However, from the study of 318 outbreaks in China, in only one the role of outdoor environmental transmission was noted. [37]

In conclusion, in the era of relaxing social distancing measures, it is important to keep the effective reproduction number below 1. Close monitoring of R_t_ is essential in order to adapt interventions over time. Its estimation is based on routine surveillance data and the delay between infection and diagnosis, resulting from the incubation time and reporting delays, can be considerable. Measuring social mixing patterns as well as adherence to infection control measures through repeated surveys can be an additional tool for real-time monitoring of the epidemic potential in the months to come.

## Materials and Methods

### Estimation of R_0_

Estimation of R_0_ was based on the number of confirmed cases with infection onset dates before the first social distancing measures were adopted (up to March 9^th^, 2020) using a maximum likelihood method and accounting for imported cases. [38] We have used the daily number of cases by date of symptom onset and inferred infection dates assuming that infection occurred approximately 5 days ago (mean incubation period [3, 39]). We assumed a gamma distributed serial interval with mean (SD) 6·67 (4·85) days. [32-34] As a sensitivity analysis, we estimated R_0_ assuming a shorter serial interval (SI Appendix, Figure S2). [36]

### A SEIR model to simulate the epidemic in Greece (February-April 2020)

We simulated the outbreak of SARS-CoV-2 in Greece from the beginning of local transmission until April 26, 2020. We assumed that local transmission initiated on February 15^th^ as the earliest reported date of symptom onset among locally infected cases was February 20^th^.

A Susceptible-Exposed-Infectious-Recovered (SEIR) model was used to simulate the outbreak (Figure 6). The population is divided into six compartments: Susceptible (S), Exposed (E), Infectious before developing symptoms (I*_pre_*), Infectious and clinically ill (I*_symp_*), true asymptomatic and partially infectious (I*_asymp_)* and Recovered (R). We assumed that a proportion *p*=80% of infected cases will develop symptoms [40] and that infectiousness may occur 1·5 days before the onset of symptoms (Table 2). [41-43] To estimate the effective reproduction number R_t_ in the presence of social distancing measures, we assessed the reduction in R_0_ using social contacts data collected from a survey implemented during lockdown (described in the next section). The impact of measures was modelled by multiplying the infection rate β by a parameter *δ* corresponding to the reduction of R_t_ in two major periods of measures: the period of initial measures including closure of schools and entertainment venues and shops (expect from super markets, groceries and pharmacies) until the day before lockdown (11 March-22 March) and the period of lockdown (23 March-26 April). More information about the model is provided in the SI Appendix.

**Figure 6.**
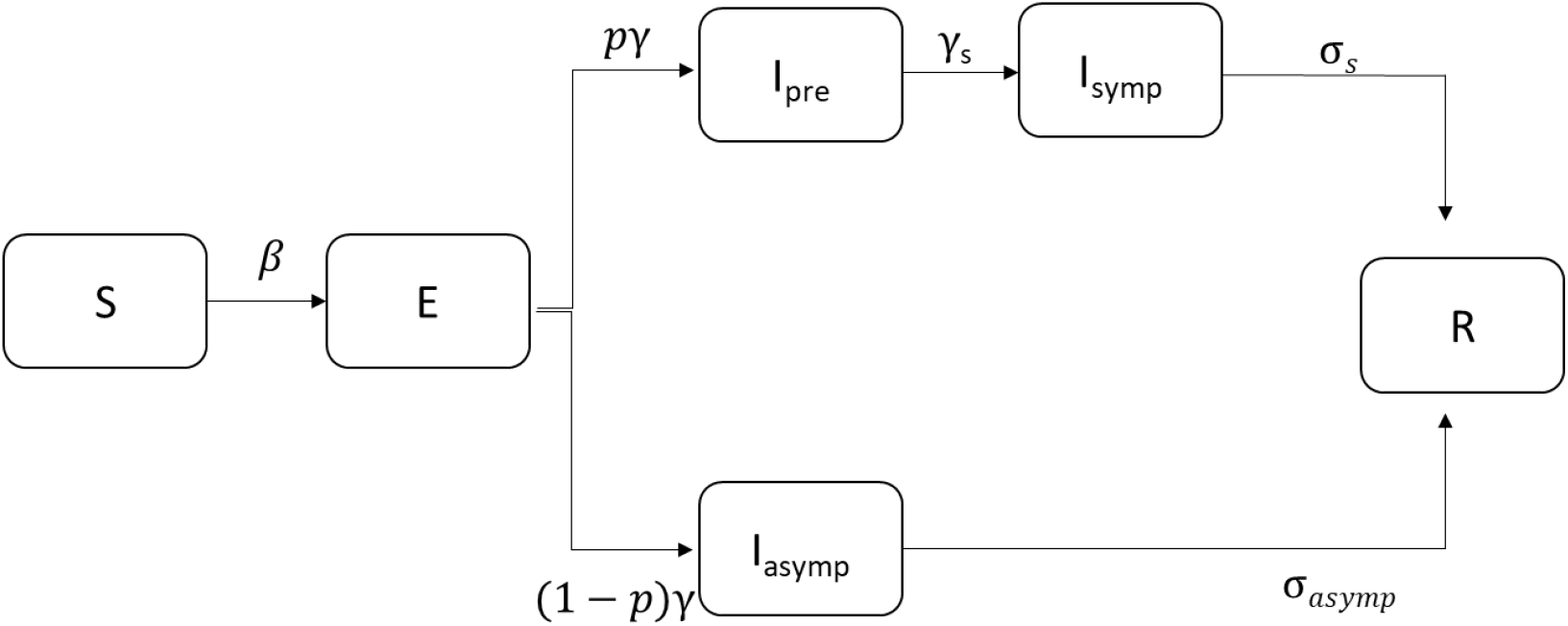
Modified SEIR model. Individuals are classified into Susceptible (S), Exposed (I), Infectious (before developing symptoms: I_pre_, clinically ill: I_symp_ or true asymptomatic: I_asymp_) and Recovered (R) states. We assumed that a proportion *p* of exposed cases will develop symptoms and that infectiousness may occur before the onset of symptoms (see also Table 1).

**Table 2.**
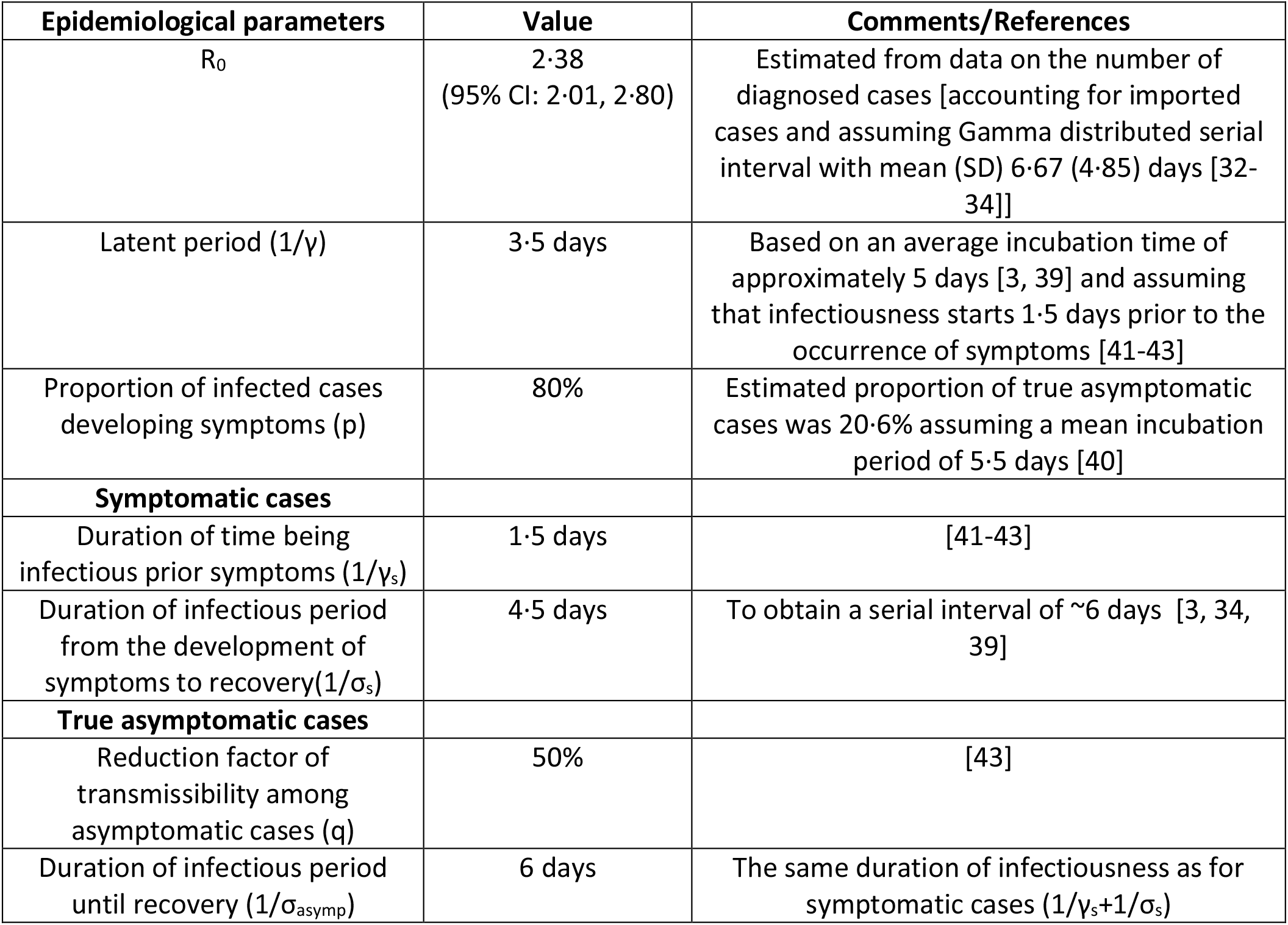
Parameters of the Susceptible-Exposed-Infectious-Recovered model.

To simulate the outbreak, we used the estimated R_0_ based on diagnosed cases in the country, other epidemiological parameters from the literature (Table 2) and the estimated reduction in transmission resulting from social distancing measures. One symptomatic case was seeded to the population at day 0 (February 15^th^) and the epidemic was further seeded by 700 imported cases over the first 40 days. This assumption was based on the number of approximately 500 imported cases diagnosed by April 7^th^ in Greece. [44] We incorporated uncertainty in R_0_ by drawing R_0_ values uniformly from the estimated 95% CI (2·01, 2·80). To account for the uncertainty in the reduction of R_0_, *δ* was drawn from a normal distribution with mean (SD) of 42·7% (1·7%) and 81·0% (1·6%) for the period of initial measures and lock-down, respectively, based on the social contacts data. One thousand simulations were performed and median estimates as well as 95% credible intervals (95% CrI) were obtained.

We obtained the infection fatality ratio (IFR) and the cumulative proportion of critically ill patients by dividing the reported number of deaths and of critically ill patients [11] to the total number of cases predicted by the model using a lag of 18 and 14 days, respectively (this lag was based on unpublished data from hospitalised patients in Greece). As patients who die on any given day are infected earlier, we have used this time lag so that the denominator reflects the total number of patients infected at the same time as those who died. [45]

To validate our findings, we used a reverse approach; we applied a published estimate of the IFR [5] to the number of infections predicted by the model and compared the resulting number of deaths to the observed (assuming again a lag of 18 days between infection and death). The published IFR estimate was adjusted to account for hon-homogeneous attack rates across age-groups, as proposed elsewhere, [12] and for the age distribution of the Greek population (SI Appendix, Table S3).

### Assessing the impact of social distancing measures

A social contacts phone survey was implemented during the period of lockdown (March 31-April 7, 2020) (SI Appendix, Figure S3). Aim of the survey was to estimate the number of contacts as well as the age-mixing of the population on a weekday during the lockdown and on the same day of the week before the pandemic (mid-January 2020) using contact diaries. In line with the POLYMOD study [46], a contact was defined as either a skin-to-skin contact (physical contact, e.g. handshake) or a two-way conversation with three or more words in the physical presence of another person (nonphysical contact). For each contact, information on the age of the person and location this contact took place (home, school, workplace, transportation, leisure, other) was recorded. We planned to recruit 600 people of all ages residing in Athens using proportional quota sampling and oversampling in the group aged between 0 and 17 years old. Participants provided oral informed consent to the interviewers. Parental-proxy completion was used for all children 0-11 years old as well as for children/adolescents 12-17 years old if the parent did not consent to provide information on their own (SI Appendix).

We assessed the impact of social distancing measures on R_t_ through the social contact matrices obtained before and during lockdown. More specifically, we defined 6 age groups (0-4, 5-11, 12-17, 18-29, 30-64, 65+) to build age-specific contact matrices before the pandemic (C*_pre_*) and during lockdown (C*_during_)* adjusting for the age distribution of the Greek population (package “socialmixr” in R). The elements c*_ij_* of these matrices denote the average number of contacts between individuals in age group *i* with individuals in age group *j*. The next-generation matrix K=(*κ_ij_*) can be parameterised using the social contact matrix as follows:

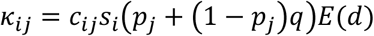

where *i,j*=1,..6 denote the age group of the participants and their contacts, respectively, *p_j_* and (1-*p_j_* is the proportion of symptomatic and true asymptomatic cases in individuals in age group *j, q* is the relative infectiousness of asymptomatic cases and *E(d)* is the average duration of infectiousness (assumed to be similar in symptomatic and asymptomatic cases). We introduce an age-dependent proportionality factor *S_i_* measuring susceptibility to infection of individuals in age group *i*. From contact tracing on clusters of confirmed cases, children are found less susceptible to infection compared to adults (SI Appendix, Table S4). We performed the analysis using a conservative estimate, i.e. that the susceptibility to infection among individuals 0-17 years old is 0·34 compared to adults. [18]

As R_0_ is the largest eigenvalue of the next generation matrix *K*, to determine its relative change during lockdown *(R_0,during_)* compared to the period before measures (R*_0,pre_)* we calculated the following ratio [47]:

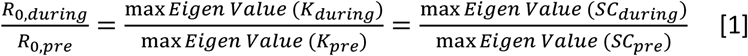

where S is a diagonal matrix accounting for the age-dependent susceptibility to infection with elements equal to 0·34 for children (*i*=1,2,3) and 1 for adults (*i*=4,5,6). A nonparametric bootstrap on the contact data by participant was performed to estimate 95% CrI for the relative change in R_0_ (n=1,000 bootstrap samples).

As multiple social distancing measures were implemented simultaneously, to disentangle the impact of each measure on R_0_ we used the information from the contacts reported on a regular weekday (January 2020) and “mimicked” the impact of each intervention separately by excluding or reducing subsets of the corresponding social contacts data (SI Appendix). [47, 48] As a contact with a particular person can take place in multiple settings (e.g. school and leisure), we assigned contacts at multiple locations to a single location using the following hierarchical order: home, work, school, leisure activities, transportation and other locations. [48] Then, we applied equation [1] using the social contacts matrix before the pandemic *(C_pre_)* and the “synthetic” matrix corresponding to each measure (or combination of measures). Using the above methodology, we also assessed scenarios with less disruptive social distancing measures as well as scenarios assessing the increase in R_t_ when measures are partially lifted after lockdown. For the scenarios after lifting lockdown, we assumed a reduction in susceptibility to infection in all age groups of 5%-30%, as a result of intensive infection control measures post lockdown (hand hygiene, use of masks, keeping distances) (SI Appendix).

## Data Availability

COVID-19 surveillance data are available through the daily reports of the Hellenic National Public Health Organization (in Greek)

https://eody.gov.gr/epidimiologika-statistika-dedomena/imerisies-ektheseis-covid-19/

## Acknowledgments

The authors would like to thank the personnel of the National Public Health Organization performing the epidemiological surveillance of SARS-CoV-2 in Greece.

The social contacts survey was funded by the Hellenic Scientific Society for the Study of AIDS and STDs.

## Conflict of interest

The authors declare that they have no conflict of interest related to this work

## Author Contributions

VS designed and programmed the model. VS and AH conceived the social contacts survey, VS, DP and AH designed the questionnaire and the protocol for the survey, VS and SR analyzed the social contacts data. AH and ST advised on the measures implemented during the epidemic in Greece. All authors reviewed the literature, advised on the parameterization of the model and the evaluated scenarios, interpreted the results, contributed to writing the article, and approved the final version for submission.

## Notes

### Competing Interest Statement

The authors have declared no competing interest.

### Author Declarations

IRB of the Hellenic Scientific Society for the Study of AIDS and STDs

## References

1. Wu F, Zhao S, Yu B, Chen YM, Wang W, Song ZG, et al. A new coronavirus associated with human respiratory disease in China. Nature 2020,579:265–269.

2. WHO. Coronavirus disease 2019 (COVID-19). Situation Report –97.

3. Li Q, Guan X, Wu P, Wang X, Zhou L, Tong Y, et al. Early Transmission Dynamics in Wuhan, China, of Novel Coronavirus-Infected Pneumonia. N Engl J Med 2020.

4. Kucharski AJ, Russell TW, Diamond C, Liu Y, Edmunds J, Funk S, et al. Early dynamics of transmission and control of COVID-19: a mathematical modelling study. Lancet Infect Dis 2020.

5. Verity R, Okell LC, Dorigatti I, Winskill P, Whittaker C, Imai N, et al. Estimates of the severity of coronavirus disease 2019: a model-based analysis. Lancet Infect Dis 2020.

6. Wu JT, Leung K, Bushman M, Kishore N, Niehus R, de Salazar PM, et al. Estimating clinical severity of COVID-19 from the transmission dynamics in Wuhan, China. Nat Med 2020,26:506–510.

7. Leung K, Wu JT, Liu D, Leung GM. First-wave COVID-19 transmissibility and severity in China outside Hubei after control measures, and second-wave scenario planning: a modelling impact assessment. Lancet 2020,395:1382–1393.

8. Greece Has an Elderly Population and a Fragile Economy. How Has It Escaped the Worst of the Coronavirus So Far? Time, April 21, 2002. https://time.com/5824836/greece-coronavirus/.

9. Tian H, Liu Y, Li Y, Wu CH, Chen B, Kraemer MUG, et al. An investigation of transmission control measures during the first 50 days of the COVID-19 epidemic in China. Science 2020.

10. Zhang J, Litvinova M, Wang W, Wang Y, Deng X, Chen X, et al. Evolving epidemiology and transmission dynamics of coronavirus disease 2019 outside Hubei province, China: a descriptive and modelling study. Lancet Infect Dis 2020.

11. National Public Health Organisation. Epidemiological surveillance of COVID-19 - Daily report. 26 April 2020 (in Greek).

12. Ferguson NM, Laydon D, Nedjati-Gilani G, Imai N, Ainslie K, Baguelin M, et al. Ferguson, N. et al. Impact of non-pharmaceutical interventions (NPIs) to reduce COVID-19 mortality and healthcare demand. Imperial College London (16–03-2020), doi:https://doi.org/10.25561/77482.

13. Imperial College London. COVID-19 model. https://mrc-ide.github.io/covid19estimates/#/details/Greece

14. Arons MM, Hatfield KM, Reddy SC, Kimball A, James A, Jacobs JR, et al. Presymptomatic SARS-CoV-2 Infections and Transmission in a Skilled Nursing Facility. N Engl J Med 2020.

15. Kimball A, Hatfield KM, Arons M, James A, Taylor J, Spicer K, et al. Asymptomatic and Presymptomatic SARS-CoV-2 Infections in Residents of a Long-Term Care Skilled Nursing Facility - King County, Washington, March 2020. MMWR Morb Mortal Wkly Rep 2020,69:377–381.

16. Park SY, Kim YM, Yi S, Lee S, Na BJ, Kim CB, et al. Coronavirus Disease Outbreak in Call Center, South Korea. Emerg Infect Dis 2020,26.

17. McMichael TM, Currie DW, Clark S, Pogosjans S, Kay M, Schwartz NG, et al. Epidemiology of Covid-19 in a Long-Term Care Facility in King County, Washington. N Engl J Med 2020.

18. Zhang J, Litvinova M, Liang Y, Wang Y, Wang W, Zhao S, et al. Changes in contact patterns shape the dynamics of the COVID-19 outbreak in China. Science 2020.

19. Jarvis CI, Van Zandvoort K, Gimma A, Prem K, group CC-w, Klepac P, et al. Quantifying the impact of physical distance measures on the transmission of COVID-19 in the UK. BMC Med 2020,18:124.

20. Prem K, Liu Y, Russell TW, Kucharski AJ, Eggo RM, Davies N, et al. The effect of control strategies to reduce social mixing on outcomes of the COVID-19 epidemic in Wuhan, China: a modelling study. Lancet Public Health 2020.

21. Davies NG, Kucharski AJ, Eggo RM, Gimma A, Edmunds WJ. The effect of non-pharmaceutical interventions on COVID-19 cases, deaths and demand for hospital services in the UK: a modelling study. *medRxiv* 2020:2020.2004.2001.20049908.

22. Pan A, Liu L, Wang C, Guo H, Hao X, Wang Q, et al. Association of Public Health Interventions With the Epidemiology of the COVID-19 Outbreak in Wuhan, China. JAMA 2020.

23. Salje H, Tran Kiem C, Lefrancq N, Courtejoie N, Bosetti P. Estimating the burden of SARS-CoV-2 in France. Pasteur-02548181. 2020.

24. Jarvis CI, Van Zandvoort K, Gimma A, Prem K, Klepac P, Rubin GJ, et al. Quantifying the impact of physical distance measures on the transmission of COVID-19 in the UK. *medRxiv* 2020:2020.2003.2031.20049023.

25. Mizumoto K, Omori R, Nishiura H. Age specificity of cases and attack rate of novel coronavirus disease (COVID-19). *medRxiv* 2020:2020.2003.2009.20033142.

26. Jing Q-L, Liu M-J, Yuan J, Zhang Z-B, Zhang A-R, Dean NE, et al. Household Secondary Attack Rate of COVID-19 and Associated Determinants. *medRxiv* 2020:2020.2004.2011.20056010.

27. Li W, Zhang B, Lu J, Liu S, Chang Z, Cao P, et al. The characteristics of household transmission of COVID-19. Clin Infect Dis 2020.

28. Danis K, Epaulard O, Benet T, Gaymard A, Campoy S, Bothelo-Nevers E, et al. Cluster of coronavirus disease 2019 (Covid-19) in the French Alps, 2020. Clin Infect Dis 2020.

29. Viner RM, Russell SJ, Croker H, Packer J, Ward J, Stansfield C, et al. School closure and management practices during coronavirus outbreaks including COVID-19: a rapid systematic review. Lancet Child Adolesc Health 2020.

30. Hoang T, Coletti P, Melegaro A, Wallinga J, Grijalva CG, Edmunds JW, et al. A Systematic Review of Social Contact Surveys to Inform Transmission Models of Close-contact Infections. Epidemiology 2019,30:723–736.

31. Zhang J, Litvinova M, Liang Y, Wang Y, Wang W, Zhao S, et al. Age profile of susceptibility, mixing, and social distancing shape the dynamics of the novel coronavirus disease 2019 outbreak in China. *medRxiv* 2020:2020.2003.2019.20039107.

32. Cereda D, Tirani D, Rovida D, Demicheli V, Ajelli M, Poletti P, et al. The early phase of the COVID-19 outbreak in Lombardy, Italy. In: *arXiv e-prints;* 2020. pp. arXiv:2003.09320.

33. Bi Q, Wu Y, Mei S, Ye C, Zou X, Zhang Z, et al. Epidemiology and Transmission of COVID-19 in Shenzhen China: Analysis of 391 cases and 1,286 of their close contacts. *medRxiv* 2020:2020.2003.2003.20028423.

34. Lavezzo E, Franchin E, Ciavarella C, Cuomo-Dannenburg G, Barzon L, Del Vecchio C, et al. Suppression of COVID-19 outbreak in the municipality of Vo, Italy. *medRxiv* 2020:2020.2004.2017.20053157.

35. Giordano G, Blanchini F, Bruno R, Colaneri P, Di Filippo A, Di Matteo A, et al. Modelling the COVID-19 epidemic and implementation of population-wide interventions in Italy. Nat Med 2020.

36. Nishiura H, Linton NM, Akhmetzhanov AR. Serial interval of novel coronavirus (COVID-19) infections. Int J Infect Dis 2020,93:284–286.

37. Qian H, Miao T, Liu L, Zheng X, Luo D, Li Y. Indoor transmission of SARS-CoV-2. *medRxiv* 2020:2020.2004.2004.20053058.

38. White LF, Wallinga J, Finelli L, Reed C, Riley S, Lipsitch M, et al. Estimation of the reproductive number and the serial interval in early phase of the 2009 influenza A/H1N1 pandemic in the USA. Influenza Other Respir Viruses 2009,3:267–276.

39. Lauer SA, Grantz KH, Bi Q, Jones FK, Zheng Q, Meredith HR, et al. The Incubation Period of Coronavirus Disease 2019 (COVID-19) From Publicly Reported Confirmed Cases: Estimation and Application. Ann Intern Med 2020.

40. Mizumoto K, Kagaya K, Zarebski A, Chowell G. Estimating the asymptomatic proportion of coronavirus disease 2019 (COVID-19) cases on board the Diamond Princess cruise ship, Yokohama, Japan, 2020. Euro Surveill 2020,25.

41. He X, Lau EHY, Wu P, Deng X, Wang J, Hao X, et al. Temporal dynamics in viral shedding and transmissibility of COVID-19. Nat Med 2020.

42. Ganyani T, Kremer C, Chen D, Torneri A, Faes C, Wallinga J, et al. Estimating the generation interval for COVID-19 based on symptom onset data. *medRxiv* 2020:2020.2003.2005.20031815.

43. Li R, Pei S, Chen B, Song Y, Zhang T, Yang W, et al. Substantial undocumented infection facilitates the rapid dissemination of novel coronavirus (SARS-CoV2). Science 2020.

44. National Public Health Organisation. Epidemiological surveillance of COVID-19 - Daily report. 7 April 2020 (in Greek).

45. Baud D, Qi X, Nielsen-Saines K, Musso D, Pomar L, Favre G. Real estimates of mortality following COVID-19 infection. Lancet Infect Dis 2020.

46. Mossong J, Hens N, Jit M, Beutels P, Auranen K, Mikolajczyk R, et al. Social contacts and mixing patterns relevant to the spread of infectious diseases. PLoS Med 2008,5:e74.

47. Hens N, Ayele GM, Goeyvaerts N, Aerts M, Mossong J, Edmunds JW, et al. Estimating the impact of school closure on social mixing behaviour and the transmission of close contact infections in eight European countries. BMC Infect Dis 2009,9:187.

48. Willem L, Hoang TV, Funk S, Coletti P, Beutels P, Hens N. SOCRATES: An online tool leveraging a social contact data sharing initiative to assess mitigation strategies for COVID-19. *medRxiv* 2020:2020.2003.2003.20030627.

